# Hard to lose, easy to gain; Trends in obesity and weight change across the life course in four British Birth cohorts

**DOI:** 10.64898/2026.06.22.26356252

**Authors:** Charis Bridger Staatz, Laura Gimeno, Demelza Smeeth, Naveed Sattar, Nish Chaturvedi, George B. Ploubidis

## Abstract

**Background:** As global obesity rates have increased, so too have efforts to manage obesity. This work estimates how many people maintain a lifelong healthy weight, have ‘weight loss potential’, and who successfully lose weight without medical support.

**Methods:** Using the 1946 National Survey of Health and Development (1946NSHD; n=4,423), the 1958 National Child Development Study (1958NCDS, n=16,749) and the 1970 British Cohort Study (1970BCS; n=15,612), we quantify the prevalence of lifetime healthy weight, overweight and obesity by ages 51-55, and compare prevalence in early adulthood to the 2000-02 Millenium Cohort Study (2001MCS; n=9,675). We identified those who maintained, lost, gained, or cycled weight up to ages 50-55 (1958NCDS) and 46-54 (1970BCS), relative to their highest body mass index (BMI) before age 42, and examined predictors of group membership using multinomial regression models.

**Findings:** In 1970BCS one-in-five people maintained a healthy BMI into their fifties, whilst 43% experienced obesity at least once, up from 25% in 1946NSHD. In 2001MCS 19% already had obesity by age 23, compared to 1-2% in the oldest cohorts. Across cohorts, those who maintained a healthy BMI were more socioeconomically advantaged, while those who experienced obesity were the most disadvantaged. Among those with obesity, a similar proportion lost weight in both cohorts (∼13%), whilst 33-39% continued weight gain. Few potential drivers were associated with weight loss after adjusting for peak BMI, whilst socioeconomic disadvantage predicted further weight gain, as did the intention to lose weight.

**Interpretation:** Weight loss from obesity is rare and the rate has remained consistent over time, whilst weight gain into obesity is common, and prevalence of lifetime obesity has increased.

**Funding:** BRC UCLH, ESRC

**Research in context:** *Evidence before this study:* A systematic review of evidence on generational differences in health from the British birth cohort studies showed that more recently born cohorts were more likely to be overweight or living with obesity than their predecessors at the same age, that they experienced an earlier onset of overweight or obesity, and that they were more likely to experience rapid increases in BMI across the life course (Seach terms were for six UK cohort studies from 1946 through to 2000-02, identifying pairwise combinations of the six cohort studies and for terms indicating the inclusion of two or more British birth cohorts. Included papers were those comparing health, including obesity, in two or more studies). While intervention studies and randomised controlled trials provide valuable evidence on short-term weight loss including its predictors, often within clinical populations, there is limited understanding of how healthy weight maintenance and sustained weight loss occur at a nationally representative, population level over longer time periods.

*Added value of this study:* This work highlights the increasing size of the population who could potentially benefit from weight loss across successive cohorts, with only 19% of people in their early 50s today having never experienced overweight or obesity. Comparatively, the same number continue to gain weight, either into or within, obesity in their early 50s. We also show that those individuals who would be most likely to benefit from weight loss tend to be the most socioeconomically disadvantaged, and that this socioeconomic gradient has remained deeply entrenched across cohorts. Our findings highlight the rarity of sustained weight loss from obesity in midlife in the era before obesity management medications were widely available, when looking at the full cohort (2.6-3.8%), and it remains uncommon even when restricting to only those with obesity at their highest BMI before there early 40s (13.1-13.7%). Any increase in the proportion of people who lost weight from obesity across cohorts, appears to be driven by the greater proportion of cohort members living with obesity rather than an increase in the propensity of those with obesity to lose weight. The majority of those with obesity and overweight peak BMIs did not just maintain overweight or obesity but continued to gain significant amounts of weight (33-39% who experienced obesity by age 42 continued to gain >5% of their bodyweight). Few indicators we tested were associated with weight loss, once peak weight was accounted for. The reported intention to lose weight was associated with continued weight gain, rather than weight loss, in fully adjusted models. Whilst there are likely other factors or unobserved drivers of weight loss not included in our analysis, one of the strongest drivers we identified of losing weight, is gaining large amounts of weight in the first place. Comparatively, being in disadvantages socioeconomic circumstances, at multiple different points across life, was related to obesity and weight gain in later mid-life.

*Implications of all the available evidence:* Despite more public health policies to help manage obesity rates at the population level, there appears to be little change in the number of people who successfully lose weight once they have developed obesity, whilst large numbers – over a third – continue to gain weight. Importantly, the intention to lose weight had little bearing on successful weight loss, suggesting that for the majority of people, will-power alone (and by implication interventions and policy that are high-agency driven), is inadequate to achieve successful weight loss. Looking forward, it appears that a large and growing proportion of the population will experience obesity at some point across adulthood - in keeping with a progressively more obesogenic environment - and therefore more may benefit from weight-loss treatment. While the emergence of GLP-1 medications is likely to disrupt historic trends, the scale of need suggests that potential demand (and costs) will be considerable, without continued efforts on preventing the development of obesity in the first place.

## Introduction

Some of the highest obesity rates in Europe are found in the UK [1], where 64% of adults in England were estimated to be overweight or had obesity in 2022-2023 [2]. More recently born cohorts experience earlier occurrence of overweight and obesity [3]. In cohorts born since the 1980s, recognised as the emergence of obesogenic environments [4], the likelihood of being overweight or developing obesity by age 10 was two to three-fold higher than earlier born cohorts [3]. Concurrently, efforts to reduce levels of obesity have increased through public health policies [5], although these have typically been high agency driven policies [6]. Theoretically, this may have positively impacted the number of people who manage to lose, maintain and gain weight across generations, but this is understudied.

Body mass index (BMI) tends to increase with age, before declining in old age [7]. Before then, downward movement across weight categories is uncommon. If weight is lost, it is often regained within a few years, known as “weight cycling”. The roll-out of newer obesity management medications (OMM; based on incretin therapies) has the potential to support effective weight loss across the population. Whilst impressive weight loss (e.g., 15-25% of body weight in one year) is seen among patients using OMMs [8], health benefits are widely reported with a more modest reduction of >5% bodyweight [9–11]. However, little is known about the pre-OMM era prevalence of sustained weight loss in the general population, or how this has changed over time. Understanding who already achieves weight loss and who continues to gain weight, and under what circumstances, is essential for anticipating the impacts of OMMs, including their implications for future population health, health-care service demand, and health inequalities.

The aim of this study was to estimate the prevalence, and socioeconomic characteristics of, people who enter adulthood at a healthy weight and remain so throughout, and those who make up the population with “weight loss potential” in four birth cohort studies, representative of people born in Britain in 1946, 1958, 1970 and 2000-02. The work leveraged the repeated measures of BMI available in the 1958 and 1970 cohorts to identify individuals who achieve sustained weight loss in later midlife, and those who gained further weight after their early forties. Given limited existing evidence on the characteristics of individuals who successfully lose weight in the general population, and those who continue to gain weight, socioeconomic, demographic and health predictors of group membership were explored.

## Methods

### Data

The 1946 National Survey of Health and Development (1946NSHD), 1958 National Child Development Study (1958NCDS), 1970 British Cohort Study (1970BCS), and 2000-2002 Millennium Cohort Study (2001MCS) are nationally representative birth cohort studies that have followed people born in England, Wales and Scotland (and Northern Ireland for 2001MCS) [12–15]. These studies have collected rich data across the life course from birth, including repeated measures of height and weight from which BMI can be calculated.

### Age- and cohort-specific prevalence of healthy, overweight and obesity

The prevalence of ‘healthy weight’ (BMI ≤ 25 kg/m^2^), overweight (BMI > 25 and ≤ 30 kg/m^2^) and obesity (BMI > 30 kg/m2) at ages 20, 23, 26 and 23 in 1946NSHD, 1958NCDS, 1970BCS and 2001MCS was described in each cohort, respectively (all measures based on self-reported weight and height). For 1946NSHD (ages 26, 36, 43 and 53, all nurse measured (NM) with exception age 26), 1958NCDS (ages 33 (NM), 42, 50 and 55), and 1970BCS (ages 30, 34, 42, 46-48 (NM) and 51-54), we described the proportion of cohort members who had always been a healthy weight, ever been overweight, or ever had obesity from their early 20s up to and including each sweep.

### Construction of weight change groups

To quantify the extent of weight gain and sustained weight loss, data was used from 1958NCDS and 1970BCS, where two a further two BMI observations were available between their mid-40s and mid-50s, to observe weight change. Peak BMI was defined as the highest observed BMI between ages 23/26 (1958NCDS/1970BCS) and 42. Sustained weight loss occurring between ages 50 and 55 in 1958NCDS, and ages 46-48 and 51-54 in 1970BCS, was defined as:

*> 5% of body weight lost from their peak BMI, sustained across two time points spanning an average of 5 years or more*.

The weight loss group was split by individuals whose peak BMI indicates obesity (>30kg/m^2^) or overweight (> 25 and ≤ 30 kg/m^2^). Individuals who maintained a healthy weight, overweight, or obesity; individuals who gained >5% of body weight within or into overweight or obesity; and individuals who weight cycled in and out, or within obesity, were also identified. These categories were also created based on BMI at 42 instead of peak BMI, to allow investigation of adult determinants of weight change. Further details are provided in the Appendix.

The prevalence of the weight change groups were estimated across the full cohort, as well as restricted to those with obesity or overweight at their peak BMI, to explore cohort differences in the *rate* of weight loss or gain among those with the ‘potential for weight loss’.

### Socioeconomic and sex composition

In 1946NSHD, 1958NCDS and 1970BCS, the composition of the ‘always healthy’, ‘ever overweight’, and ‘ever had obesity’ groups were examined according to parental social class and the cohort members educational attainment. Advantaged childhood social class was defined as having a father who worked in a professional or managerial profession (up to age 15 in 1946NSHD, and at birth in 1958NCDS and 1970BCS), and degree-level education was defined as having National Vocational Qualifications (NVQ) level ≥4 (by age 43 in 1946NSHD, 30 in 1958NCDS and 33 in 1970BCS).

### Predictors of selection into weight change groups

To explore drivers of selection into weight change groups in 1958NCDS and 1970BCS, data was extracted on a range of characteristics across the life course: Parental occupational social class (age 0); childhood housing tenure (age 5/7); birthweight (age 0); breastfeeding (measured retrospectively at age 5/7); childhood BMI and presence of medical conditions (age 10/11); adolescent psychological distress (age 16); educational attainment and equivalised household income in adulthood (age 30/33); smoking, partnership status and longstanding illness (age 42); and in 1970BCS only, a measure indicating intention to lose weight (age 42). Further description of the variables are given in the Appendix.

For each hypothesised driver in turn, multinomial logistic regression models were constructed, with weight change groups as the outcome and ‘maintains healthy weight’ as the reference category. For potential adult drivers of weight change, models used the weight change outcome derived relative to BMI at age 42 rather than peak. This ensures appropriate temporal ordering between determinants and subsequent weight change. We ran models adjusting for predictor-specific confounders selected a priori using Directed Acyclic Graphs (DAGs) (see Appendix). For both child and adult determinants, we run models adjusting for peak BMI, therefore identifying predictors of subsequent weight change, rather than achieved highest weight. For adult models, this adjustment accounts for drivers of selection into the weight change groups. Model outputs were presented as average marginal effects (AMEs), which reflect the percentage-point difference in the predicted probability of being in each weight change group for those with and without each predictor. We primarily focused our reporting on the weight loss from obesity and weight gain into or within obesity groups, with full results for the other obesity experienced groups in the Appendix.

### Missing data

To mitigate the impact of nonresponse and attrition on our results [16–19], we used multiple imputation by chained equations (MICE; in 1946NSHD, 1958NCDS and 1970BCS) generating minimum 15 imputed datasets for each study, imputing data to the target population (those alive and living in Britain at age 53 in 1946NSHD, 51–54 in 1970BCS and 55 in 1958NCDS). For 2001MCS we use inverse probability weights for nonresponse. More information is provided in the Appendix.

## Results

### Healthy weight, overweight and obesity across the life course

Already in cohort members’ early-mid 20s, there were clear cohort differences (Figure 1). Whilst 85.91% [95% CI: 84.54, 87.27] of 1946NSHD and 80.84% [95% CI: 80.13, 81.56] of 1958NCDS had a BMI <25 kg/m^2^ this was only the case for 66.10% [95% CI: 64.98, 67.23] of 1970BCS, dropping further to 54.11% [95% CI: 52.48, 55.72] of 2001MCS at age 23. The proportion of cohort members who were overweight or living with obesity in their early 20s was therefore substantially higher in later cohorts. For instance, 19.42% [95% CI: 18.10, 20.83] of 2001MCS had obesity at age 23, compared to 6.47% [95% CI: 6.02, 6.91] of 1970BCS at age 26, 2.43% [95% CI: 2.17, 2.69] of 1958NCDS at age 23, and 1.09% [95% CI: 0.72, 1.47] of 1946NSHD at age 20.

**Figure 1.**
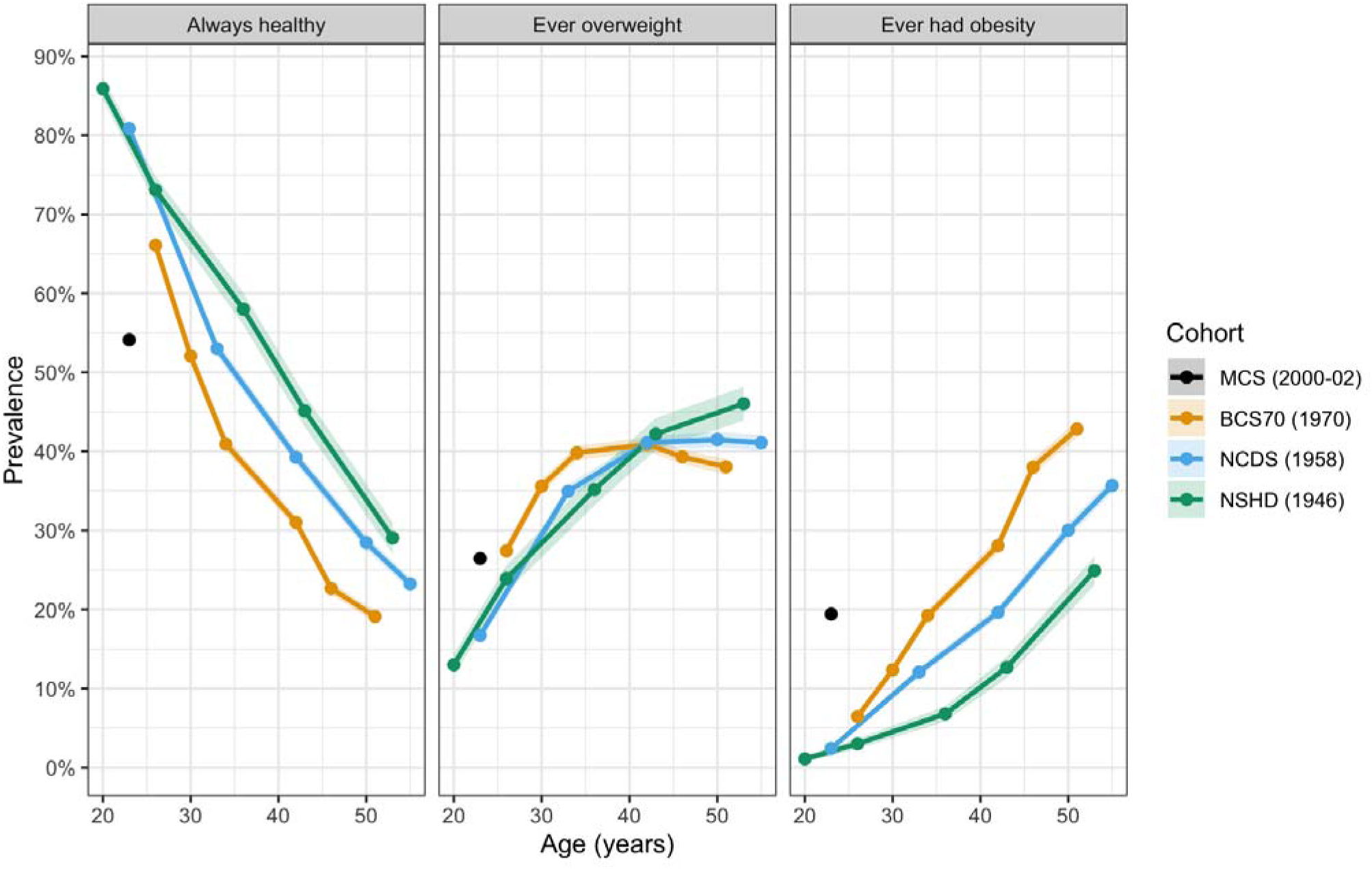
Prevalence of ‘always healthy,’ ‘ever overweight,’ ‘ever had obesity’ in four British birth cohorts. Prevalence of life-time healthy, overweight and obesity. In 2001MCS, 1970BCS, 1958NCDS majority of height and weight measures are self-reported, except at age 46-48 in 1970BCS and 33 in 1958NCDS. In NSHD, the majority of height and weight measures, with the exception of age 20 and 26, were objectively collected. Note that the overweight groups sees cumulative prevalence decline, as it is the only groups people can both move into (from always healthy) and out of (into ever had obesity).

The proportion of cohort members who successfully maintained a healthy weight between their early 20s and early to mid-50s declined dramatically, from approximately 29.06% [95% CI: 27.07, 31.04] of 1946NSHD to 19.11% [95% CI: 18.37, 19.84] of 1970BCS cohort members. This decline appeared to be compensated primarily by an increase in the proportion of cohort members who ever had obesity, which increased from 24.90% [95% CI: 23.07, 26.74] in 1946NSHD, to 35.69% [95% CI: 34.75, 36.63] in 1958NCDS and 42.83% [95% CI: 41.76, 43.91] in 1970BCS, such that in the 1970BCS, ‘ever had obesity’ was the largest group at age 51. The age at which the ‘always healthy’ group became the minority shifted to progressively younger ages.

Individuals in the ‘always healthy’ group in their early to mid-50s tended to be more socioeconomically advantaged than the cohort average (Figure 2) in all three cohorts. They were more likely to have a father who worked in a professional or managerial occupation, and to have post-secondary qualifications. On the other hand, the socioeconomic profile of those in the ‘ever had obesity’ group tended to be less advantaged than the cohort average.

**Figure 2.**
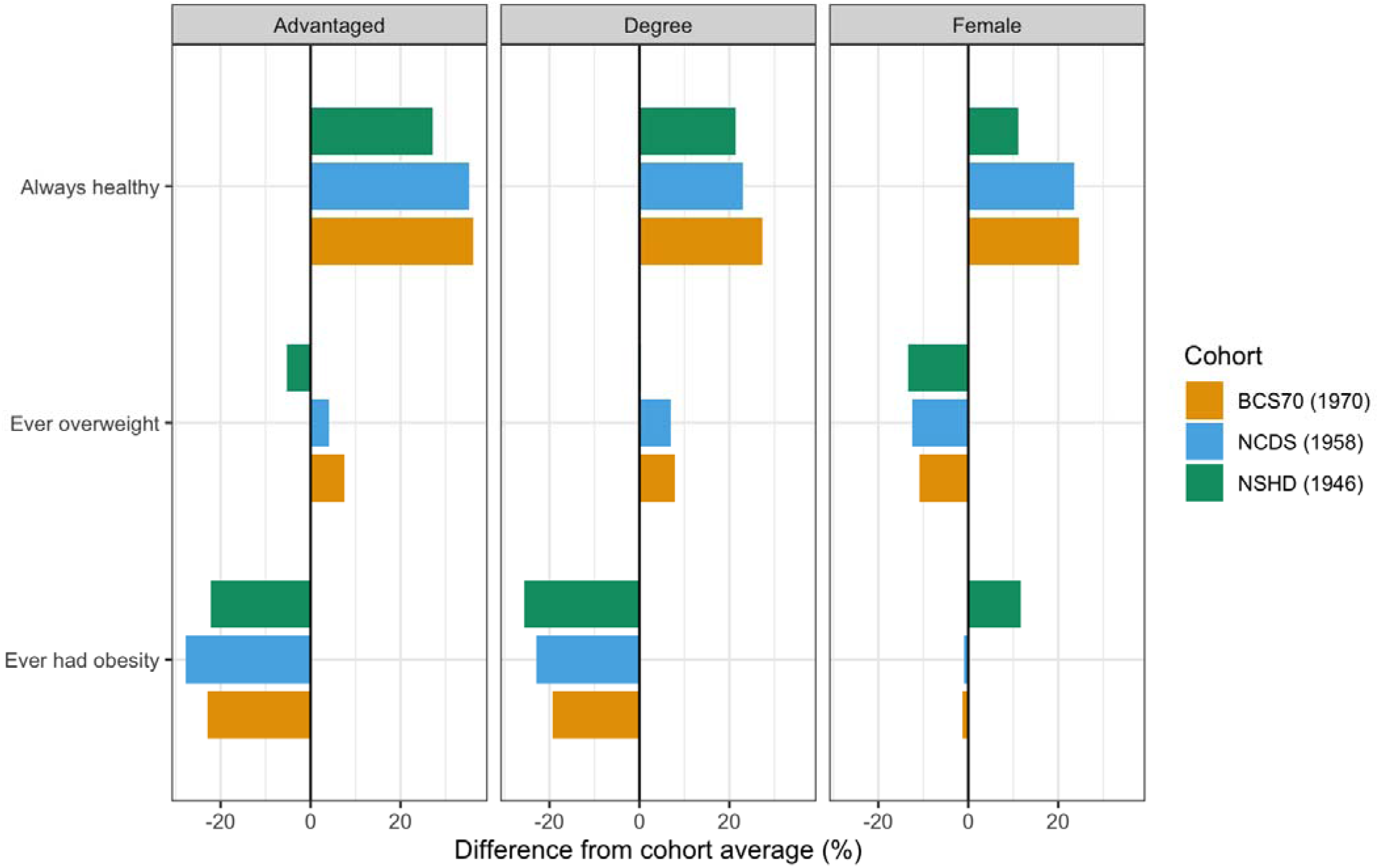
Socioeconomic and sex composition of the ‘always healthy,’ ‘ever overweight’ and ‘ever had obesity’ groups by early to mid-50s in three British birth cohorts. Advantaged social class refers to cohort members with parents who worked in professional or managerial occupations (Registrar General Social Class I or II). Degree refers to cohort members with a National Vocational Qualification (NVQ) level ≥ 4. All comparisons are made relative to the cohort average, to take into consideration changes in the distribution of occupation and education across cohorts.

### Prevalence of weight loss and weight gain

In both cohorts, only a small percentage of the overall cohort lost >5% weight from obesity (2.57% [95% CI: 2.29, 2.86] in 1958NCDS, 3.84% [95% CI: 3.40, 4.29] in 1970BCS) or from overweight (4.29% [95% CI: 3.81, 4.77] in 1958NCDS, 5.72% [95% CI: 5.18, 6.26] in 1970BCS) (Figure 3). Nearly one-in-five of the overall cohort continued to gain weight within or into obesity (1958NCDS 19.21% [95% CI: 18.40, 20.02]; 1970BCS 18.78% [95% CI: 17.78, 19.78]).

**Figure 3.**
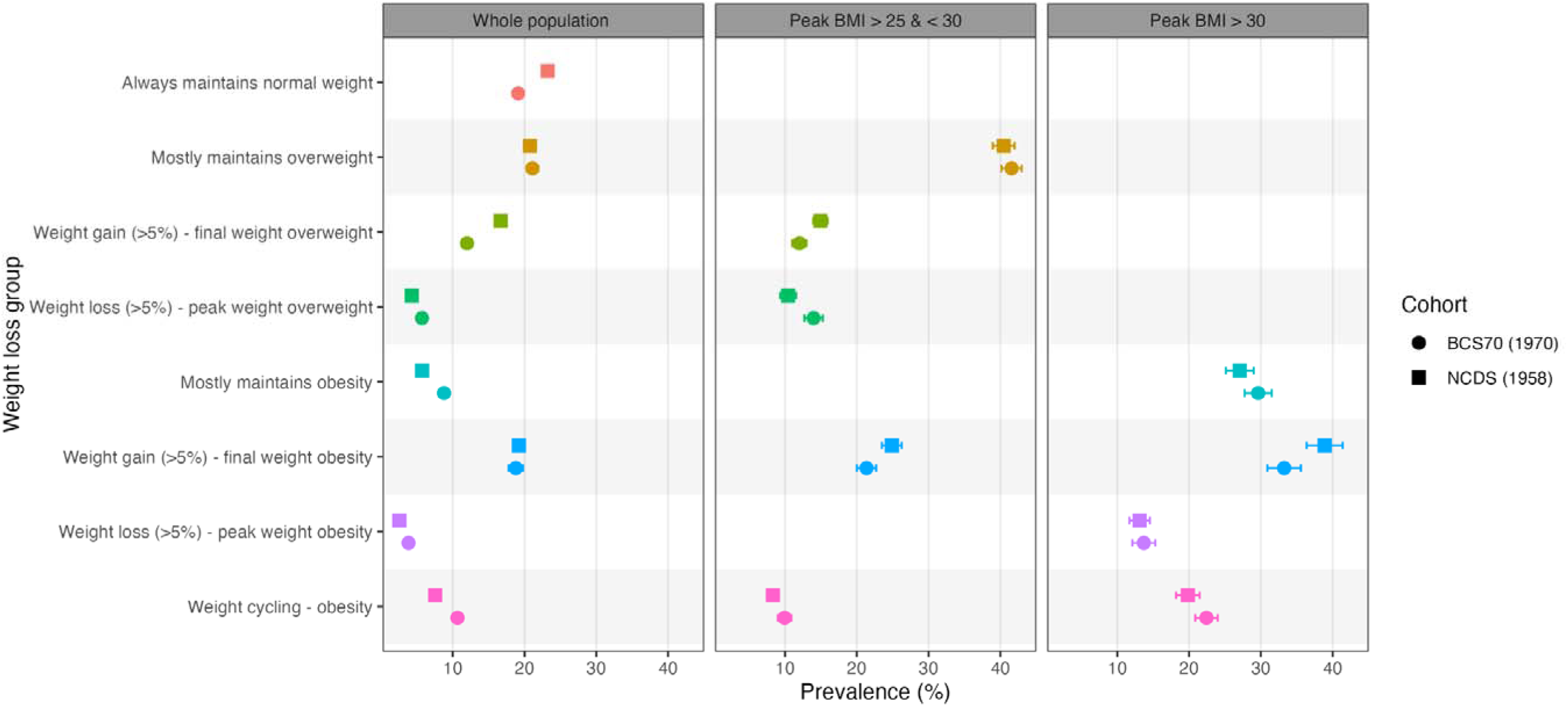
Prevalence of weight change and maintenance groups in the 1958 and 1970 cohort at ages 46-55, including restricted to those with prior overweight and obesity. Panel 1 shows the proportion of weight change and maintenance groups in the full cohort sample, whilst panel 2 restricts to those whose peak BMI was classified as overweight (>25kg/m^2^ and ≤ 30kg/m^2^) and panel 3 among those whose peak BMI indicates obesity (>30kg/m^2^). Weight change was defined between ages of 50-55 in NCDS, and 46-54 in BCS70, relative to the highest BMI (‘peak’) observed between the ages of 23/26 and 42.

Considering only those whose peak BMI was >30kg/m^2^, and therefore could potentially lose weight from obesity, a similarly modest proportion of cohort members lost weight and maintained weight loss in both cohorts (1958NCDS: 13.11% [95% CI: 11.69, 14.53]; 1970BCS: 13.69% [95% CI: 12.10, 15.29]). More than one third continued to gain weight (38.88% [95% CI: 36.37, 41.39] in 1958NCDS and 33.24% [95% CI: 30.92, 35.56] in 1970BCS). Amongst those with an overweight peak BMI, 13.99% [95% CI: 12.70, 15.27] lost weight in 1970BCS and 10.43% [95% CI: 9.33, 11.54] in 1958NCDS. A greater proportion of people who had overweight peak BMIs went on to gain further weight, either within overweight (14.93 [95% CI: 13.92, 15.94] in 1958NCDS and 12.00 [95% CI: 11.00, 12.99] in 1970BCS) or into obesity (24.87 [95% CI: 23.50, 26.24] in 1958NCDS and 21.36 [95% CI: 20.02, 22.70] in 1970BCS).

### Predictors of selection into weight change groups

In both 1970BCS and 1958NCDS, before adjusting for achieved peak BMI, a higher childhood BMI increased the likelihood of losing weight from obesity in their late 50s (Figure 4, Appendix Table A9 & A10. 1958NCDS AME=0.007, 95% CI: 0.006 to 0.008, p < 0.001; 1970BCS AME= 0.010, 95% CI: 0.008 to 0.012, p < 0.001). In 1970BCS, being male (AME = −0.010, 95% CI: −0.018 to −0.002, p=0.020), in a disadvantaged social class at birth (Class IV/V AME = 0.018, 95% CI: 0.007 to 0.028, p < 0.001), and being in rented housing (AME= 0.013, 95% CI: 0.002 to 0.023, p= 0.020) also increased the likelihood of weight loss from obesity, whilst in 1958NCDS being female was the only variable that also increased this likelihood (AME= 0.006, 95% CI: 0.001 to 0.012, p= 0.016). However, after adjusting for peak BMI, none of the explored drivers remain associated with weight loss from obesity in either cohort, with the exception of being male in 1970BCS, suggesting these early life factors increase risk of developing obesity, rather than subsequent weight loss.

**Figure 4.**
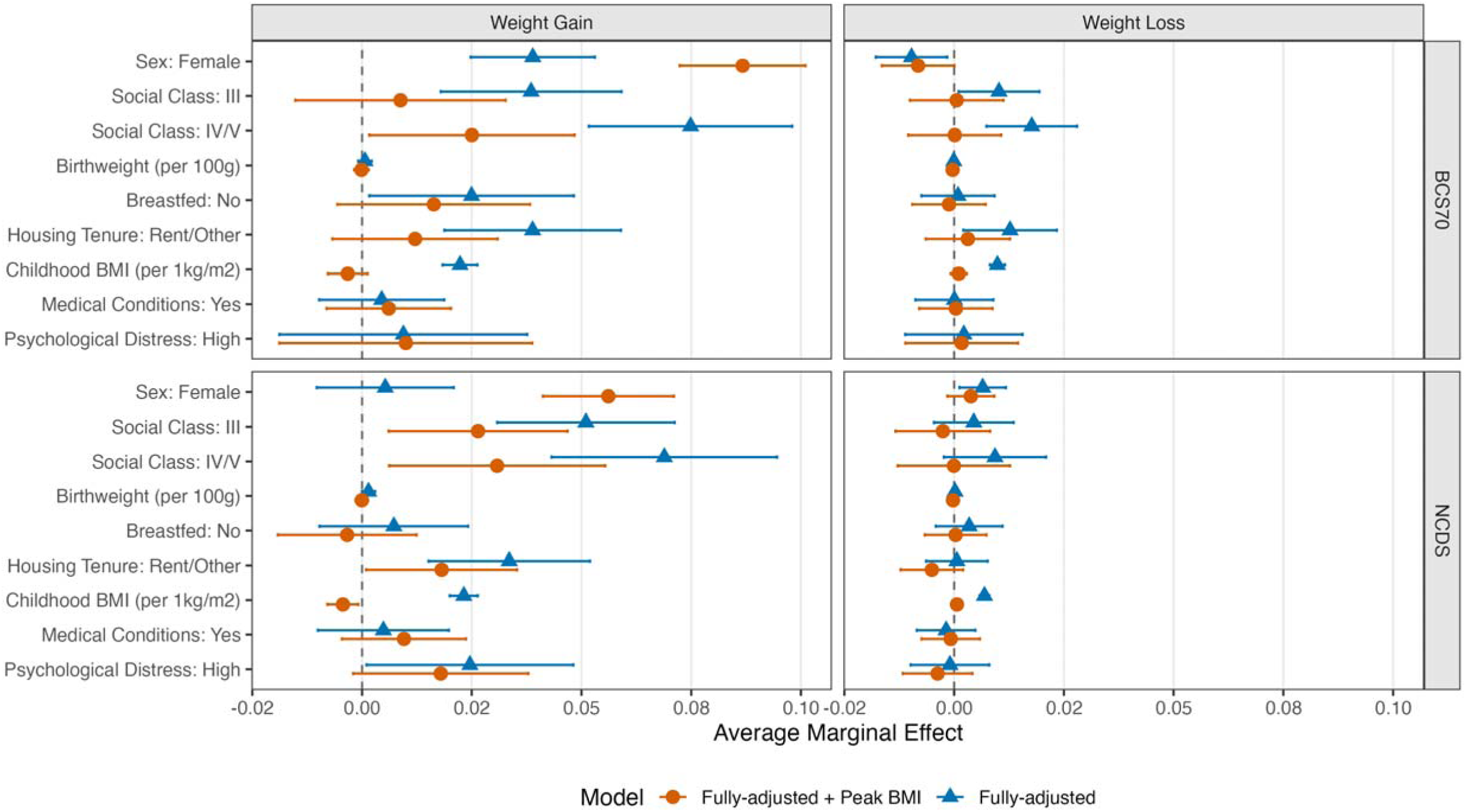
Early life determinants of selection into obesity weight gain and obesity weight loss groups. For each individual driver of weight change, separate multinomial logistic regressions analysis were run, adjusting for a unique set of covariates, as deemed appropriate for the specific driver, using Directed Acyclic Graphs (DAGs) as described in the Appendix. For each of the above models, the reference groups are as follows: Sex – male; Social Class - class I/II (managerial, professional, technical); Breast feeding – yes; Housing Tenure – own outright or with a mortgage; Medical Conditions – none; Psychological Distress – low.

In 1970BCS, after adjusting for peak BMI, only being in the most disadvantaged social class at birth remained related to continued weight gain into or within obesity (Figure 4, Table A9: Social Class I/IV AME= 0.025, 95% CI: 0.002 to 0.048, p = 0.036). In 1958NCDS, before adjustment for peak BMI (Figure 4, Table A10), disadvantaged social class at birth (Social Class IV/V AME= 0.069, 95% CI: 0.043 to 0.095, p < 0.001), higher birthweight (AME= 0.001, 95% CI: 0.000 to 0.003, p = 0.052), living in rented housing in childhood (AME= 0.034, 95% CI: 0.015 to 0.052, p < 0.001) and experiencing adolescent psychological distress (AME= 0.025, 95% CI: 0.001 to 0.048, p = 0.041) predict later life weight gain within or into obesity, in addition to higher childhood BMI (AME= 0.023, 95% CI: 0.020 to 0.026, p < 0.001). These associations largely remained after adjustment for peak BMI, except associations with birthweight were fully attenuated, and the association with childhood BMI changed direction (AME= −0.004, 95% CI: −0.008 to −0.001, p = 0.016). In other words, assuming individuals reach the same peak BMI, those who had a lower BMI in childhood and who therefore gained weight more rapidly in adulthood, were at greater risk of continued weight gain in their 50s.

When looking at potential drivers of weight change in adulthood (exploring change and maintenance relative to BMI at 42, rather than peak BMI. See Appendix for proportions in each group), and conditioning on peak BMI, none of the explored drivers increased the likelihood of later life weight loss (Figure 5, Appendix Table A11 and A12). This included the intention to try and lose weight, measured in 1970BCS only, which was only related to subsequent weight loss prior to controlling for peak BMI. Comparatively, the intention to lose weight was associated with subsequent weight gain into or within obesity, even after adjusting for peak BMI (AME= 0.02, 95% CI: 0.01 to 0.04, p = 0.010). Additionally, being in the lowest income quintile and not having a university degree was associated with subsequent weight gain into or within obesity in 1970BCS (lowest income: AME= 0.03, 95% CI: 0.01 to 0.06, p = 0.008; No degree: AME= 0.02, 95% CI: 0.00 to 0.04, p = 0.021). In both cohorts, being married or in a civil partnership reduced the risk of weight gain (1958NCDS: AME= −0.02, 95% CI: −0.04 to 0.00, p = 0.026; 1970BCS: AME= −0.02, 95% CI: −0.03 to 0.00, p = 0.020).

**Figure 5.**
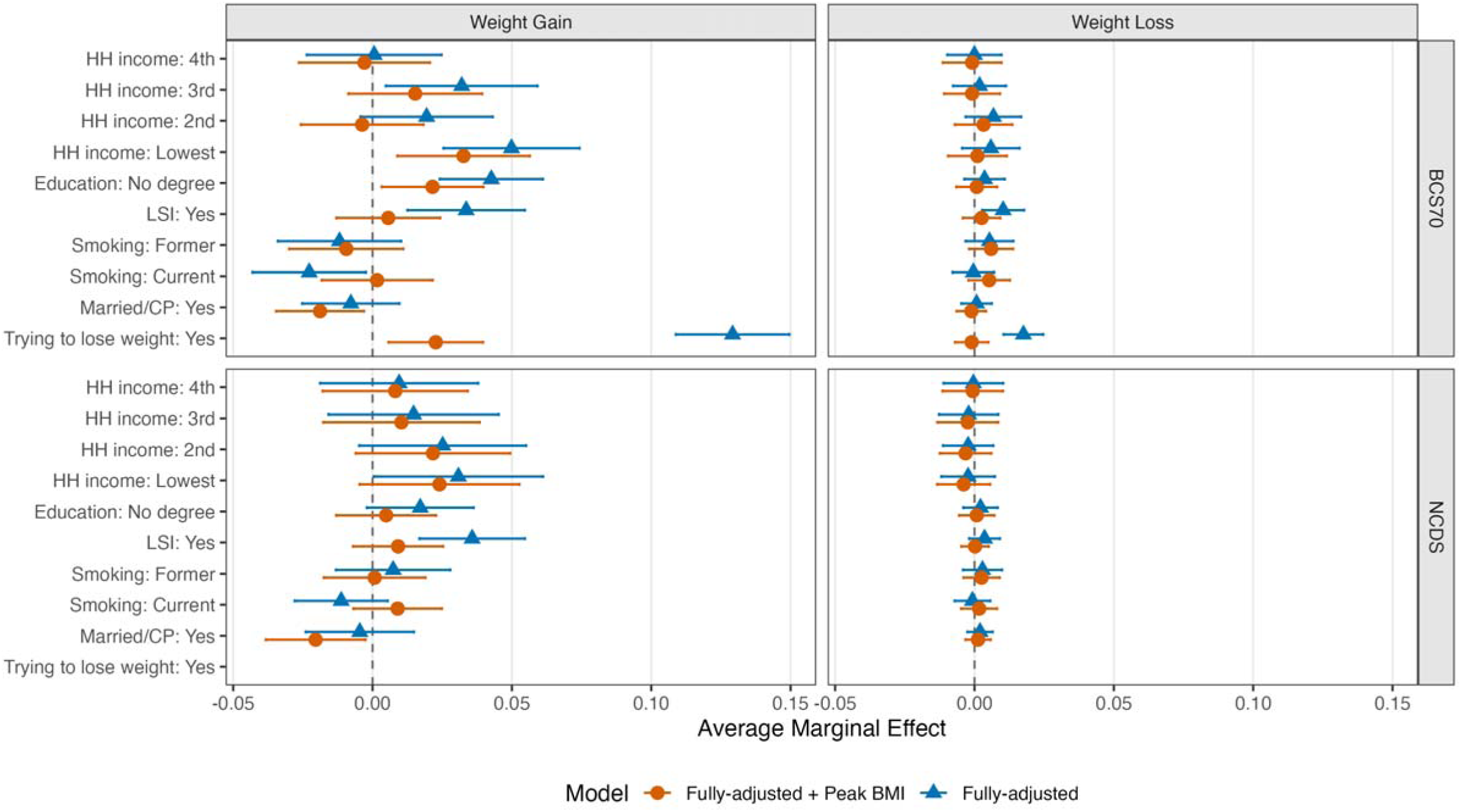
Adult determinants of selection into obesity weight gain and obesity weight loss groups. Figure 5 uses a measure of weight change defined against BMI at age 42, rather than based on peak BMI, and makes further adjustment for peak BMI as a driver of selection into the groups. For each individual driver of weight change, separate multinomial logistic regressions analysis were run, adjusting for a unique set of covariates, as deemed appropriate for the specific driver, using Directed Acyclic Graphs (DAGs) as described in the Appendix. For each of the above models, the reference groups are as follows: Income – highest; Education: - has a degree; Long Standing Illness (LSI) – none; Partnership Status – not legally married (or in a civil partnership); Trying to lose weight – no (measured in BCS70 only).

## Discussion

The lifetime prevalence of overweight and obesity has increased in younger generations. Consequently, the proportion of people who consistently maintain a healthy weight throughout adulthood has decreased dramatically, to fewer than one-in-five among people in their early 50s today. Already at age 23, nearly half of those born in 2000-02 were overweight or living with obesity, a striking rise compared to earlier generations. Our findings highlight the sheer increase in the number of people who might benefit from weight loss, who also tend to be more socioeconomically disadvantaged, and therefore may face greater challenges in doing so. This study also shows that in an era before widespread availability of OMM, losing weight once obesity has emerged, and sustaining that weight loss, is difficult: less than one-in-seven achieved weight loss. Comparatively, it remains easy to continue gaining weight: over a third of individuals with obesity by their early 40s continued to gain weight in their 50s. The challenge of resisting an obesogenic environment is simply too hard to overcome for many [4, 20], so that ∼87% simply do not lose meaningful weight once obesity manifests.

The number of people losing weight from obesity in the total cohort was marginally higher in 1970BCS than in 1958NCDS. Comparisons of child to adolescent BMI transitions in 1958NCDS and 2001MCS have similarly found that the odds of being ‘overweight decreasing to normal weight’ were higher in the younger cohort [21]. However, this should not be read uncritically. Our study suggests that such trends are symptomatic of the higher prevalence of obesity in the population (i.e., larger proportion with obesity who could potentially lose weight) and reflects a regression from high levels of BMI, rather than a shift toward healthier weight norms. Once stratifying by overweight and obesity peak BMIs, we also observed typically higher prevalence of later life weight gain in 1958NCDS than in 1970BCS. This likely reflects that 1970BCS experienced increasingly obesogenic environments across childhood and adolescence, whilst later life weight gain in 1958NCDS likely reflects a delayed, life-stage–specific response to increasingly obesogenic environments kicking in around the 1980s [4]. Relatedly, early life determinants of weight gain remain predictive in 1958NCDS after adjustment for peak BMI, but not in 1970BCS. Later exposure to obesogenic environments in 1958NCDS likely drove weight gain among those with high susceptibility to obesity but who’s early environments did not promote weight gain.

Existing research on predictors of weight loss, summarised in a systematic review, has come primarily from experimental trials conducted in the United States, with shorter follow-up times than our study [22]. The review found little evidence of demographic determinants of weight loss and only found evidence of behavioural and cognitive determinants. Our study provides a valuable life course perspective on this question. The early life predictors we identified of weight loss from obesity, notably higher childhood BMI and disadvantaged SEP, were fully attenuated after adjustment for peak BMI, suggesting much of the association is actually with obesity rather than subsequent weight loss. We found that among individuals with obesity in 1970BCS, intention to lose weight had little bearing on eventual weight loss but was instead related to continued weight gain. Evidence from the cross-sectional National Health and Nutrition Examination Surveys in the United States found that the desire to lose weight is more common in higher BMI categories [23]. In our study, individuals with obesity who intended to lose weight were more likely to gain weight, highlighting that lack of willpower is unlikely driving current trends in obesity [4, 20].

A major strength of this study is its use of large, nationally representative prospective cohort studies which have followed people born in different generations since birth [16–18, 24]. This means that repeated measures of BMI could be used to identify weight change or maintenance groups, and that a diverse set of predictors of group membership could be considered, including early life factors. Despite loss to follow-up, the richness of the data collected and known properties of the in the British Birth Cohorts at baseline mean that approaches to missing data, including MICE as used in this study, are effective in restoring representativeness to the target population [16–19].

Our approach to characterising weight loss differs from latent class and related trajectory modelling approaches [25, 26], which only occasionally reveal clear or robust classes characterised by weight loss [21]. Weight change groups were defined using fixed percentage and BMI cut-offs, which will inevitably simplify individual variation in weight trajectories. As a result, some individuals may have been misclassified, particularly those who weight cycled between observations. Most weight and height measures were self-reported in 1970BCS and 1958NCDS, whilst the majority of measures in 1946NSHD were objective nurse measures. Self-reported weight is typically underestimated in women, and height overestimated in men [27], and therefore the cohort differences presented likely underestimate the true differences. Although we attempt to account for unintended weight loss due to illness, by examining long-standing illness as a driver of weight change, it remains possible that unobserved health factors continue to influence the weight changes observed, and there remains the possibility of unmeasured confounding on the selection into weight change groups in the analysis.

Our analyses present patterns of weight change in a pre-OMM era. Future prevalence of weight loss will increase with the expansion of OMMs, and the sociodemographic predictors of weight loss group membership will likely change, particularly since early adopters of OMM may be more socioeconomically advantaged. An estimated 3% of adults were using OMMs in Britain in early 2025, with the majority of prescriptions obtained privately [28]. Analysis of private prescription medication shows that fewer individuals from deprived areas use OMM once accounting for higher rates of obesity, and they start using them at a higher BMI level indicating advanced health risk [29]. Wider availability and use of OMMs may also have implications for the size and composition of the weight cycling group.

In the future, a large and growing proportion of the population will experience obesity at some point across adulthood and could therefore benefit from weight-loss treatment. Our work shows that in a pre-OMM are only a small proportion of individuals go on to lose weight, indicating the challenge in achieving weight loss once obesity has developed, whilst a large proportion continue to gain weight with significant implications for health. These cohort trends, which show lower levels of obesity are possible, point to significant changes in the environment over the 20^th^ century that have promoted modern patterns of obesity. Worryingly, the rates of obesity seen in the youngest generation in their early 20s are similar to the proportion who ever experienced obesity by their late 40s in the 1946 cohort, with likely significant consequences for their future health. While OMMs will likely disrupt historic obesity trends, and be important in future obesity strategy, the scale of need for these medications will be high without simultaneous focus on tackling the increasingly obesogenic environments.

## Funding

This work was supported by the UK Economic and Social Research Council which provides core funding for the UCL Centre for Longitudinal Studies), and the National Child Development Study (NCDS), 1970 British Cohort Study (BCS70), and Millennium Cohort Study (BCS70) (grant number ES/W013142/1). The UK Medical Research Council provides core funding for the Unit for Lifelong Health and Ageing at UCL and the MRC National Survey of Health and Development (grant numbers MC-UU-0019/1).

This research was also supported by the Economic and Social Research Council (ES/S012583 and ES/W013142/1) and the National Institute for Health Research University College London Hospitals Biomedical Research Centre (BRC1007/MMI/GP/101420) and was carried out in collaboration with NHS England.

## Supporting information

Paper Appendix

## Data Availability

The data used in this study from the National Child Development Study (NCDS), 1970 British Cohort Study (BCS70), and Millennium Cohort Study (BCS70) are available from the UK Data Service. Access to these data is subject to registration and may require approval from the CLS Data Access Committee to ensure the confidentiality of participants. Further information on accessing the CLS data can be found on the UK Data Service website: https://ukdataservice.ac.uk/. Data from the MRC National Survey of Health and Development (NSHD) are curated by the Unit for Lifelong Health and Ageing at UCL and are available to bona fide researchers via application through the NSHD data-sharing platform (Skylark: www.skylark.ucl.ac.uk/NSHD). An NSHD dataset is also deposited with the UK Data Service (DOI: 10.5255/UKDA-SN-8732-3).

## Acknowledgements

This research and the future research these data will enable, would not be possible without the valuable contributions of the NSHD, NCDS, BCS70 and MCS cohort members over many years. We are very grateful for their ongoing commitment to the studies.

## Declaration of Interest (Conflicts of Interest)

CBS, LG, DS, and GBP have no conflict of interests to declare. NC receives funds from AstraZeneca to serve on Data Safety and Monitoring Committees for clinical trials. N.S. has consulted for and/or received speaker honoraria from AbbVie, Amgen, AstraZeneca, Boehringer Ingelheim, Carmot Therapeutics, Eli Lilly, Gan & Lee, GlaxoSmithKline, Hanmi Pharmaceuticals, Kailera Therapeutics, Mass Medicines, Menarini Ricerche, Metsera, Novo Nordisk, Pfizer, Regeneron, Roche, UCB Pharma, and Verdiva Bio, and received grant support paid to his university from AstraZeneca, Boehringer Ingelheim, Novartis, and Roche outside the submitted work.

## Data Sharing Statement

The data used in this study from the National Child Development Study (NCDS), 1970 British Cohort Study (BCS70), and Millennium Cohort Study (BCS70) are available from the UK Data Service. Access to these data is subject to registration and may require approval from the CLS Data Access Committee to ensure the confidentiality of participants. Further information on accessing the CLS data can be found on the UK Data Service website: https://ukdataservice.ac.uk/.

Data from the MRC National Survey of Health and Development (NSHD) are curated by the Unit for Lifelong Health and Ageing at UCL and are available to bona fide researchers via application through the NSHD data-sharing platform (Skylark: www.skylark.ucl.ac.uk/NSHD). An NSHD dataset is also deposited with the UK Data Service (DOI: 10.5255/UKDA-SN-8732-3).

## Author Contributions

CBS developed the initial idea for the paper. CBS and LG analysed and interpreted the data, LG created all figures, CBS and LG drafted the manuscript, and all authors (DS, NS, NC, GBP) critically revised the manuscript for important intellectual content and gave final approval for the version published.

## Notes

### Author Declarations

This study used de-identified data from four British birth cohort studies. The relevant Research Ethics Committees of the institutions responsible for each cohort gave ethical approval for the original data collection and subsequent waves of follow-up. Participants provided informed consent according to the requirements of each study. The present analyses were conducted under the relevant data access agreements and institutional approvals. The data used in this study from the National Child Development Study (NCDS), 1970 British Cohort Study (BCS70), and Millennium Cohort Study (BCS70) are available from the UK Data Service. Access to these data is subject to registration and may require approval from the CLS Data Access Committee to ensure the confidentiality of participants. Further information on accessing the CLS data can be found on the UK Data Service website: https://ukdataservice.ac.uk/. Data from the MRC National Survey of Health and Development (NSHD) are curated by the Unit for Lifelong Health and Ageing at UCL and are available to bona fide researchers via application through the NSHD data-sharing platform (Skylark: www.skylark.ucl.ac.uk/NSHD).

