## Supplementary material for "Hard to lose, easy to gain; Trends in obesity and weight change across the life course in four British Birth cohorts": Paper Appendix

### Appendix A1. Missing Data and Imputation used to derive weight change groups in BCS70 and NCDS

**Appendix Table A.1. List of auxiliary and substantive variables used in imputation model, and description of variable type.**

| **Variable** | **Description** | **BCS70: Age Measured** | **NCDS: Age Measured** |
| --- | --- | --- | --- |
| **Variables used to derive weight loss and weight trajectories** | | | |
| Weight (kg) | Continuous | 26, 30, 34, 42, 46-48, 51-54 | 23, 33, 42, 44, 50, 55 |
| Lifetime Height (m)  Derived from height at observations in adulthood | Continuous | 26, 30, 34, 42, 46-48, 51-54 | 23, 33, 42, 44, 50, 55 |
| **Socioeconomic** | | | |
| Parent read to child | Binary | 5 | 7 |
| Mothers Age Left Education | Binary | Birth | Birth |
| Fathers Age Left Education | Binary | Birth | 7 |
| Fathers Social Class | Ordinal | Birth | Birth |
| Adult Social Class / Manual Class | Ordinal | 34 | 33, 42 |
| Adult NVQ | Ordinal | 30 | 33 |
| Childhood Housing Tenure | Binary | 5 | 7 |
| Adult Housing Tenure | Binary | 42 | 42 |
| Childhood Household Income | Ordinal | 10 | 16 |
| Adult Household Income | Ordinal | 30 | 33 |
| Economic Activity | Ordinal | 42, 51-54 | 42, 55 |
| **Demographic** | | | |
| Voting in last election | Binary | 42 | 42 |
| Number of times family moved | Ordinal | 5 | 7 |
| Has someone to listen to them | Ordinal | 42 | - |
| Member of Organisation | Ordinal | 42 | 33 (binary – union membership), 42 |
| Adult Marriage Status | Binary | 42 | 42 |
| Number of rooms in house/per person | Continuous/ Ordinal | 5 | Birth |
| Country of Birth | Binary | Birth | Birth |
| Age outcome measured | Continuous | 51-54 | 55 |
| Sex at birth | Binary | Birth | Birth |
| Region in Adulthood | Unordered | 42 | 42 |
| **Health** | | | |
| Long standing Illness | Binary | 26, 30, 34, 42 | 33, 42 |
| New Cancer Diagnosis Ages 42-51 | Binary | 42-51 | 42 |
| Breastfed | Binary | 5 | 7 |
| Maternal Mental Health – Malaise | Binary | 5 | - |
| Adult Mental Health - Malaise | Binary | 34 | 33 |
| Maternal Smoking | Ordinal | Birth | Birth |
| Adult Self-rated Health | Binary | 42 | 42 |
| Smoking Status | Ordinal | 30, 42 | 33, 42 |
| Cognition (Principal Component) | Continuous | 10 | 11 |
| Birthweight | Continuous | Birth | Birth |
| Child Medical Conditions | Binary | 10 | 11 |
| Child BMI | Continuous | 10 | 11 |
| Adolescent Mental Health - Malaise | Binary | 16 | 16 |
| Adolescent BMI | Continuous | 16 | 16 |
| Desire to change weight | Binary | 42 | - |
| Self-assessment of weight | Binary | - | 42 |
| Biomarkers: Total Cholesterol, HDL Cholesterol, HbA1C, BCS70 only: Systolic Blood Pressure, Diastolic Blood Pressure  NCDS only: Insulin-like Growth Factor 1 (IGF-1), fibrinogen, C-Reactive Protein | Continuous | 46 | 44 |
| **Auxiliary Variable: Participation in the Sweep** | | | |
| Participation in the sweep | Binary | 51-54 | 55 |

### Appendix A2. Prevalence of always a normal weight, ever overweight and ever obese groups in NSHD, NCDS, BCS70 and MCS

We used data from four cohorts to describe the prevalence of always a normal weight, ever overweight and ever obese across the life course.

- NSHD (born 1946): ages 20, 26, 36, 43, and 53
- NCDS (born 1958): ages 23, 33, 42, 50 and 55
- BCS70 (born 1970): ages 26, 30, 34, 42, 46 and 51
- MCS (born 2000-02): age 23

At each age, we calculated Body Mass Index (BMI) using adult height and sweep-specific weight. We categorised cohort members at each sweep into normal/underweight (BMI < 25 kg/m2), overweight (BMI ≥ 25 and < 30 kg/m2) and obese (BMI ≥ 30 kg/m2). Having established each cohort members’ BMI category in their early 20s, we derived indicators at each subsequent sweep for whether the cohort member had *ever* experienced overweight (but not obesity) and *ever* experienced obesity up to and including that sweep.

To mitigate the risk of bias from nonresponse and attrition, we imputed data in NSHD, NCDS and BCS70 to the target population (alive and resident in the UK at the time of the final sweep, at age 53 in NSHD [n = 4423], 55 in NCDS [n = 16,749] and 51 in BCS70 [n = 15,612]). We used auxiliary variables predictive of nonresponse and of BMI itself in imputation models, taken from across the life course. These included measures of early life socioeconomic position, mental and physical health (including childhood height and BMI), health-related behaviours (e.g., smoking), and measures of social participation (e.g., voting, membership in organisations), as well as previous patterns of nonresponse. We generated minimum 15 imputed datasets for each of NSHD, NCDS and BCS70.

Prevalences for NSHD were estimated using survey weights to account for study design, which oversampled cohort members with fathers in non-manual and agricultural professions. In MCS, since no cumulative indicators were being derived, we estimated the prevalence of normal weight, overweight and obesity using data from respondents at the age 23 sweep only (n = 9,675), applying inverse probability weights for nonresponse to restore representativeness to the target population, combined with survey weights to account for the complex sampling design of MCS.

### Appendix A3. Defining weight loss groups in BCS70 and NCDS

We defined weight loss in the final two sweeps of BCS70 (age 46-48 and age 51-54) and NCDS (age 50 and 55), known as the ‘observation period’, as weight loss of greater than 5% compared to the highest recorded weight in adulthood (peak weight) observed between the ages of 23/26 and 42.

**Maintained normal weight.** Individuals who had a healthy (or underweight) peak BMI (≤25 k/m2) between ages 23/26 and 42 and maintained a healthy (or underweight) BMI during the observation period. Some individuals in this group may lose or gain weight across the life course, but their BMI remains ≤25kg/m^2^ at each observation.

**Mostly maintained overweight.** Individuals who had an overweight peak BMI (>25kg/m^2^ and ≤30kg/m^2^) between ages 23/26 and 42 and/or and maintained this weight without additional weight gain (>5% of body weight) or weight loss (>5% of body weight). This group also included individuals who lost more than 5% of body weight in the final observation (at age 51-54 in BCS70, and age 55 in NCDS) only, but for whom ‘sustained’ weight loss could not be confirmed.

This group also included individuals whose peak BMI was healthy but moved into the overweight group without substantial weight gain (>5%) in either of the final two observations, or the alternative where their weight reduced by less than 5% from peak, which may have moved them to a healthy weight. Individuals who weight cycled (see fluctuations in their weight) but experience one observation in the overweight category at either at peak BMI or in the observation window are also included in this group. This group also included a few marginal cases, where both peak and final BMI were overweight but bordering on obesity. This group didn’t experience any significant weight gain or loss (>5%), but their nurse-measured weight at 46-48 in BCS70 put them marginally in the obese category.

**Weight gain into overweight.** This group includes people who gained more than 5% of their body weight in the final observation, relative to their peak BMI before age 42, and their final BMI is consider overweight (>25kg/m^2^ and ≤30kg/m^2^)

**Weight loss from overweight.** This group includes people who ‘sustained >5% weight loss’, but from a peak BMI >25kg/m^2^ and ≤30kg/m^2^ at both time points in the observation window. This group includes both individuals whose weight loss returned their weight to healthy in the final two observations or remained in the overweight category.

**Mostly maintained obesity.** Individuals who had an obese peak BMI (>30kg/m^2^) between ages 23/26 and 42 and maintained this weight without additional weight gain (>5% of body weight) or weight loss (>5% of body weight). This group also included individuals who lost more than 5% of body weight in the final observation (at age 51-54 in BCS70, and age 55 in NCDS) only, but for whom ‘sustained’ weight loss could not be confirmed.

This group also contained a number of marginal cases, where no significant weight loss or gain occurred (>5%) between peak BMI and the observational period, but peak BMI or final BMI was obese (>30kg/m^2^).

**Obese – gain >5% of body weight.** Individuals who gained more than 5% of body weight compared to their previously highest recorded weight, and whose final BMI was >30kg/m^2^. This category could also include individuals that were previously healthy weight or overweight based on their peak BMI, but who gained weight to end up with an obese BMI at age 51-54 (BCS70) or age 55 (NCDS), as well as individuals who were already overweight or obese, and further increased their weight >5%.

**Obese – weight loss >5% of body weight.** Individuals who lost more than 5% of body weight at both observations in the final two sweeps compared to a previous obese peak BMI (>30kg/m^2^) between ages 23/26 and 42. Weight loss may occur within the obesity BMI class or take cohort members from an obese BMI to an overweight or healthy BMI. This group excludes individuals who lost >5% of body weight at only one of the two final observations.

**Obese – weight cycling.** Broadly speaking, these were individuals who either lost or gained 5% relative to their peak weight between ages 23/26 and 42, but for who this was only observed at one of the two final observations (i.e., not sustained). For these individuals, their BMI was >30kg/m^2^ at either peak, age 46-48 or 51-54 in BCS70 and age 50 or 55 in NCDS, or at multiple observations.

More specifically, this group included:

- Individuals that experienced 5% weight loss at age 46-48/50 compared to their peak weight but returned to within 5% of peak weight at the final observation, and who had an obese final BMI.
- Individuals that experienced 5% weight loss at 46-48/50 compared to their peak weight, then gained an additional 5% at age 51-54/55 compared to their peak weight, and who were obese at either peak or the final observation.
- Individuals that experienced 5% weight gain at 46-48/50, but whose BMI returned to within 5% of peak by the final observation and had an obese BMI during the observation period.
- Individuals that experienced 5% weight gain at 46-48/50, then experienced 5% weight loss in the final observation compared to their peak BMI, and had an obese BMI at either peak or the final observation.
- Individuals whose weight was 5% higher compared to peak at both 46-48/50 and 51-54/55. Notable weight gain occurred at age 46-48/50, and weight was lost relative to 46-48/50 but not peak BMI. These individuals experienced a large amount of weight gain at 46-48/50, taking them from normal/borderline overweight (peak BMI) to obese (age 46-48/50) to overweight (51-54/55).
- Individuals whose weight was 5% lower than peak at 46-48/50, having come from marginally obese peak BMI but then returning to within 5% of peak by age 51-54/55.

Given the intervals between sweeps it is likely that there are “weight cyclers” in the other groups, but we were unable to identify them with the available data.

We did not use nurse-measured BMI at age 44 in NCDS to define lifetime prevalence groups, peak BMI, or subsequent weight loss, to avoid measurement mode effect. Including nurse-measured BMI as part of the observations used to define peak BMI would risk overestimating weight loss, since subsequent BMI observations used to identify weight loss were based on self-reports, which likely underestimate true BMI. In contrast, nurse-measured BMI at age 46-48 in BCS70 was retained, since in this case it is not used to identify peak BMI but rather as one of the observations used to identify weight loss during the observation period, and is therefore a more conservative estimate of weight loss, although it may increase the number of individuals classified as “weight cyclers”.

### Appendix A4: Full Model Specification and minimal sufficient adjustment sets

**Appendix Table A2.** Description of potential drivers of weight loss, age data was collected, and the minimal sufficient adjustment sets applied in multinomial logistic regression models

| **Variable of Interest** | **Age Measured** | **Minimal sufficient adjustment sets** |
| --- | --- | --- |
| Sex  (Ref: Male; Female) | Birth | Age |
| Social occupational class (Ref: II/I Managerial and Professional; III Manual/Non-Manual; V/IV Unskilled/Partly Skilled) | Birth | Age, sex, age mother left full-time education |
| Birth weight  (per 100g) | Birth | Age, sex, age mother left full-time education, social occupational class, maternal smoking in pregnancy |
| Breastfeeding  (Ref: Yes; No) | 5/7 | Age, sex, age mother left full-time education, birthweight, social occupational class |
| Housing tenure  (Ref: Owns outright/with a mortgage; Rents or other). | 5/7 | Age, sex, age mother left full-time education, social occupational class |
| Childhood BMI  (per 1kg/m^2^) | 10/11 | Age, sex, age mother left full-time education, social occupational class, birthweight, breastfeeding, housing tenure, maternal smoking in pregnancy |
| Medical conditions  (Ref: No; Yes) | 10/11 | Age, sex, age mother left full-time education, social occupational class, birthweight, breastfeeding, housing tenure, maternal smoking in pregnancy |
| Adolescent psychological distress  (Ref: Low Malaise (9-item Malaise score < 4); High Malaise (9-item Malaise score ≥4)) | 16 | Age, Sex, age mother left full-time education, Social Occupational Class, birthweight, breast feeding, housing tenure, Childhood BMI, Medical Conditions, Childhood cognition |
| Equivalised Household Income Quintiles  (Ref: Highest Quintile) | 30/33 | Age, Sex, age mother left full-time education, Social Occupational Class, housing tenure, childhood cognition, adolescent psychological distress |
| Highest Educational Attainment (Ref: Degree (NVQ4-5); No degree (NVQ1-3)) | 30/33 | Age, Sex, age mother left full-time education, Social Occupational Class, housing tenure, childhood cognition, adolescent psychological distress |
| Smoking status  (Ref: Never; Former; Current) | 42 | Age, sex, age mother left full-time education, social occupational class, housing tenure, childhood cognition, adolescent psychological distress, education level, adult psychological distress |
| Longstanding illness  (Ref: No; Yes) | 42 | Age, sex, age mother left full-time education, social occupational class, housing tenure, childhood BMI, medical conditions, childhood cognition, adolescent psychological distress, education level, adult psychological distress |
| Partnership status  (Ref: Not married/civil partnership; Married/civil partnership) | 42 | Age, Sex, age mother left full-time education, Social Occupational Class, housing tenure, medical conditions, childhood BMI, education, childhood cognition, adolescent psychological distress, adult psychological distress |
| *1970BCS only*  Intention to lose weight  (Ref: Not trying to lose weight; Yes trying to lose weight) | 42 | Age, sex, age mother left full-time education, social occupational class, housing tenure, childhood BMI, childhood cognition, adolescent psychological distress, education, adult psychological distress, longstanding illness |

### Appendix A5: Lifetime prevalence healthy weight, overweight and obesity by cohort

**Appendix Table A3.** Lifetime healthy, overweight and obese in 1946NSHD

|  | NSHD | | |
| --- | --- | --- | --- |
| Weight Loss Groups | % | L CI | U CI |
| Age 20 – Self Reported | | | |
| Always Healthy | 85.91 | 84.54 | 87.27 |
| Ever Overweight | 13 | 11.66 | 14.34 |
| Ever Had Obesity | 1.09 | 0.72 | 1.47 |
| Age 26 – Self Reported | | | |
| Always Healthy | 73.09 | 71.33 | 74.85 |
| Ever Overweight | 23.9 | 22.19 | 25.6 |
| Ever Had Obesity | 3.01 | 2.36 | 3.67 |
| Age 36 – Nurse Measured | | | |
| Always Healthy | 58.03 | 55.98 | 60.07 |
| Ever Overweight | 35.16 | 33.14 | 37.17 |
| Ever Had Obesity | 6.81 | 5.84 | 7.79 |
| Age 43 – Nurse Measured | | | |
| Always Healthy | 45.12 | 43.07 | 47.17 |
| Ever Overweight | 42.21 | 40.19 | 44.23 |
| Ever Had Obesity | 12.67 | 11.31 | 14.03 |
| Age 53 – Nurse Measured | | | |
| Always Healthy | 29.06 | 27.07 | 31.04 |
| Ever Overweight | 46.04 | 43.9 | 48.17 |
| Ever Had Obesity | 24.90 | 23.07 | 26.74 |

**Appendix Table A4.** Lifetime healthy, overweight and obesity in 1958NCDS

|  | NCDS – Main Analysis | | | NCDS – Sensitivity with Nurse Measured BMI included | | |
| --- | --- | --- | --- | --- | --- | --- |
| Weight Loss Groups | % | L CI | U CI | % | L CI | U CI |
| Age 23 – Self Reported | | | | | | |
| Always Healthy | 80.84 | 80.13 | 81.56 | - | - | - |
| Ever Overweight | 16.73 | 16.04 | 17.42 | - | - | - |
| Ever Had Obesity | 2.43 | 2.17 | 2.69 | - | - | - |
| Age 33 – Nurse Measured | | | | | | |
| Always Healthy | 53.00 | 52.01 | 53.99 | - | - | - |
| Ever Overweight | 34.94 | 34.09 | 35.79 | - | - | - |
| Ever Had Obesity | 12.06 | 11.42 | 12.69 | - | - | - |
| Age 42 – Self Reported | | | | | | |
| Always Healthy | 39.26 | 38.32 | 40.19 | - | - | - |
| Ever Overweight | 41.11 | 40.23 | 41.96 | - | - | - |
| Ever Had Obesity | 19.63 | 18.89 | 20.38 | - | - | - |
| Age 44 - Nurse Measured (excluded from main analysis) | | | | | | |
| Always Healthy | - | - | - | 29.23 | 28.36 | 30.09 |
| Ever Overweight | - | - | - | 42.17 | 41.28 | 43.05 |
| Ever Had Obesity | - | - | - | 28.60 | 27.78 | 29.43 |
| Age 50 – Self Reported | | | | | | |
| Always Healthy | 28.47 | 27.55 | 29.34 | 24.95 | 24.08 | 25.83 |
| Ever Overweight | 41.49 | 40.54 | 42.44 | 41.62 | 40.70 | 42.54 |
| Ever Had Obesity | 30.04 | 29.14 | 30.94 | 33.43 | 32.49 | 34.37 |
| Age 55 – Self Reported | | | | | | |
| Always Healthy | 23.22 | 22.43 | 24.01 | 21.21 | 20.44 | 21.98 |
| Ever Overweight | 41.09 | 40.10 | 42.09 | 40.89 | 39.91 | 41.87 |
| Ever Had Obesity | 35.69 | 34.75 | 36.63 | 37.90 | 36.93 | 38.87 |

**Appendix Table A4 Footnote**: To limit measurement mode effect, the nurse measured BMI values at age 44 was excluded from the main analysis. As sensitivity, the age 44 nurse measured weight are included in the construction of lifetime prevalence.
**Appendix Table A5.** Lifetime healthy, overweight and obese in 1970BCS

|  | BCS70 | | |
| --- | --- | --- | --- |
| Weight Loss Groups | % | L CI | U CI |
| Age 26 – Self Reported | | | |
| Always Healthy | 66.10 | 64.98 | 67.23 |
| Ever Overweight | 27.43 | 26.43 | 28.43 |
| Ever Had Obesity | 6.47 | 6.02 | 6.91 |
| Age 30 – Self Reported | | | |
| Always Healthy | 52.06 | 51.12 | 53.00 |
| Ever Overweight | 35.59 | 34.71 | 36.48 |
| Ever Had Obesity | 12.35 | 11.73 | 12.97 |
| Age 34 – Self Reported | | | |
| Always Healthy | 40.93 | 40.00 | 41.86 |
| Ever Overweight | 39.82 | 38.90 | 40.73 |
| Ever Had Obesity | 19.25 | 18.48 | 20.01 |
| Age 42 – Self Reported | | | |
| Always Healthy | 31.02 | 30.18 | 31.86 |
| Ever Overweight | 40.90 | 40.01 | 41.79 |
| Ever Had Obesity | 28.08 | 27.11 | 29.06 |
| Age 46 – Nurse Measured | | | |
| Always Healthy | 22.67 | 21.88 | 23.46 |
| Ever Overweight | 39.31 | 38.43 | 40.20 |
| Ever Had Obesity | 38.02 | 37.01 | 39.03 |
| Age 51 – Self Reported | | | |
| Always Healthy | 19.11 | 18.37 | 19.84 |
| Ever Overweight | 38.06 | 37.07 | 39.06 |
| Ever Had Obesity | 42.83 | 41.76 | 43.91 |

**Appendix Table A6.** Lifetime healthy, overweight and obese in 2001MCS

|  | MCS – Main Results | | | MCS – Sensitivity (Non-white ethnicity removed) | | |
| --- | --- | --- | --- | --- | --- | --- |
| Weight Loss Groups | % | L CI | U CI | % | L CI | U CI |
| Age 23 – Self Reported | | | | | | |
| Always Healthy | 54.11 | 52.48 | 55.72 | 54.24 | 52.44 | 56.02 |
| Ever Overweight | 26.47 | 25.23 | 27.74 | 26.68 | 25.31 | 28.10 |
| Ever Had Obesity | 19.43 | 18.10 | 20.83 | 19.08 | 17.82 | 20.40 |

**Appendix Table A6 Footnote**: MCS is an ethnically diverse cohort, whereas the older cohorts are primarily white-British. As a sensitivity analysis, we provide prevalence of healthy weight, overweight and obesity in those of white British ethnicity only.

### Appendix A6: Proportion in weight change groups

**Appendix Table A7.** Proportion in weight change groups in BCS70 and NCDS relative to peak BMI.

|  | BCS70 | | | NCDS | | |
| --- | --- | --- | --- | --- | --- | --- |
| Weight Loss Groups | **%** | **L CI** | **U CI** | **%** | **L CI** | **U CI** |
| Always Maintains Normal Weight | 19.11 | 18.37 | 19.84 | 23.21 | 22.42 | 24.01 |
| Mostly Maintains Overweight | 21.09 | 20.29 | 21.90 | 20.75 | 19.95 | 21.56 |
| Weight gain (>5%) - final weight overweight | 12.00 | 11.31 | 12.68 | 16.67 | 15.92 | 17.42 |
| Weight loss (>5%) - peak weight overweight | 5.72 | 5.18 | 6.26 | 4.29 | 3.81 | 4.77 |
| Mostly maintains Obese | 8.79 | 8.12 | 9.46 | 5.73 | 5.28 | 6.17 |
| Weight gain (>5%) - final weight Obese | 18.78 | 17.78 | 19.78 | 19.21 | 18.40 | 20.02 |
| Weight loss (>5%)- peak weight Obese | 3.85 | 3.40 | 4.29 | 2.57 | 2.29 | 2.86 |
| Weight Cycling - Obese | 10.67 | 9.99 | 11.34 | 7.56 | 7.07 | 8.05 |
| Weight Loss Group - Peak BMI > 30 |  |  |  |  |  |  |
| Mostly Maintains Overweight | 1.01 | 0.64 | 1.38 | 1.11 | 0.63 | 1.59 |
| Mostly maintains Obese | 29.63 | 27.74 | 31.53 | 27.07 | 25.13 | 29.02 |
| Weight gain (>5%) - final weight Obese | 33.24 | 30.92 | 35.56 | 38.88 | 36.37 | 41.39 |
| Weight loss (>5%)- peak weight Obese | 13.69 | 12.10 | 15.29 | 13.11 | 11.69 | 14.53 |
| Weight Cycling - Obese | 22.43 | 20.86 | 23.99 | 19.83 | 18.18 | 21.48 |
| Weight Loss Group - Peak BMI > 25 & <30 |  |  |  |  |  |  |
| Mostly Maintains Overweight | 41.56 | 40.15 | 42.97 | 40.46 | 38.94 | 41.98 |
| Weight gain (>5%) - final weight overweight | 12.00 | 11.00 | 12.99 | 14.93 | 13.92 | 15.94 |
| Weight loss (>5%) - peak weight overweight | 13.99 | 12.70 | 15.27 | 10.43 | 9.33 | 11.54 |
| Mostly maintains Obese | 1.15 | 0.76 | 1.54 | 0.99 | 0.65 | 1.33 |
| Weight gain (>5%) - final weight Obese | 21.36 | 20.02 | 22.70 | 24.87 | 23.50 | 26.24 |
| Weight Cycling - Obese | 9.95 | 8.99 | 10.90 | 8.31 | 7.48 | 9.14 |

**Appendix Table A7 Footnote:** Due to a small number of individuals being marginal cases, and being allocated to groups as described above, there are some individuals who are grouped as “mostly maintains overweight” despite having a peak BMI indicative of obesity, and a small number grouped as “mostly maintains obesity” whose peak BMI is overweight.

**Appendix Table A8.** Proportion in weight change groups in BCS70 and NCDS relative to BMI at age 42.

|  | BCS70 | | | NCDS | | |
| --- | --- | --- | --- | --- | --- | --- |
| Weight Loss Groups | **%** | **L CI** | **U CI** | **%** | **L CI** | **U CI** |
| Always Maintains Normal Weight | 21.43 | 20.69 | 22.18 | 24.81 | 24.02 | 25.61 |
| Mostly Maintains Overweight | 19.20 | 18.24 | 20.15 | 19.19 | 18.41 | 19.97 |
| Weight gain (>5%) - final weight overweight | 15.49 | 14.71 | 16.28 | 19.27 | 18.50 | 20.05 |
| Weight loss (>5%) - peak weight overweight | 2.83 | 2.45 | 3.22 | 2.35 | 1.96 | 2.73 |
| Mostly maintains Obese | 7.46 | 6.91 | 8.00 | 4.84 | 4.38 | 5.30 |
| Weight gain (>5%) - final weight Obese | 20.81 | 19.82 | 21.80 | 20.74 | 19.90 | 21.58 |
| Weight loss (>5%)- peak weight Obese | 2.26 | 1.93 | 2.58 | 1.37 | 1.15 | 1.59 |
| Weight Cycling - Obese | 10.52 | 9.85 | 11.19 | 7.42 | 6.87 | 7.98 |

**Appendix Table A8 Footnote:** To explore adult determinants of change, and to maintain causal ordering of drivers and timing of weight change, the outcome group was derived based on BMI at 42, rather than peak BMI, to ensure that drivers occurred before the change in weight was observed.

| BCS70 Early Life Drivers of Weight Change | Obesity Weight Maintenance | | Obesity Weight Gain | | Obesity Weight Loss | | Obesity Weight Cycling | |
| --- | --- | --- | --- | --- | --- | --- | --- | --- |
|  | AME (95% CI) | P Value | AME (95% CI) | P Value | AME (95% CI) | P Value | AME (95% CI) | P Value |
| Minimally Sufficient Adjustment sets | | | | | | | | |
| Sex: Female | -0.037 (-0.048 to -0.026) | < 0.001 | 0.039 (0.025 to 0.053) | < 0.001 | -0.010 (-0.018 to -0.002) | 0.020 | 0.003 (-0.010 to 0.016) | 0.652 |
| Social Class: III | 0.020 (0.006 to 0.034) | 0.005 | 0.039 (0.018 to 0.059) | < 0.001 | 0.010 (0.001 to 0.019) | 0.030 | 0.027 (0.013 to 0.042) | < 0.001 |
| Social Class: IV/V | 0.031 (0.014 to 0.048) | < 0.001 | 0.075 (0.052 to 0.098) | < 0.001 | 0.018 (0.007 to 0.028) | < 0.001 | 0.043 (0.024 to 0.062) | < 0.001 |
| Birthweight (per 100g) | 0.001 (0.000 to 0.002) | 0.029 | 0.001 (-0.001 to 0.002) | 0.398 | 0.000 (-0.001 to 0.001) | 0.845 | 0.000 (-0.001 to 0.002) | 0.582 |
| Breastfed: No | 0.001 (-0.011 to 0.014) | 0.815 | 0.025 (0.002 to 0.048) | 0.037 | 0.001 (-0.007 to 0.009) | 0.834 | 0.005 (-0.009 to 0.018) | 0.497 |
| Housing Tenure: Rent/Other | 0.014 (0.002 to 0.026) | 0.020 | 0.039 (0.019 to 0.059) | < 0.001 | 0.013 (0.002 to 0.023) | 0.020 | 0.018 (0.004 to 0.031) | 0.010 |
| Childhood BMI (per 1kg/m^2^) | 0.020 (0.017 to 0.023) | < 0.001 | 0.022 (0.018 to 0.026) | < 0.001 | 0.010 (0.008 to 0.012) | < 0.001 | 0.017 (0.014 to 0.020) | < 0.001 |
| Medical Conditions: Yes | -0.002 (-0.015 to 0.011) | 0.763 | 0.004 (-0.010 to 0.019) | 0.538 | 0.000 (-0.009 to 0.009) | 0.990 | -0.005 (-0.017 to 0.008) | 0.468 |
| Psychological Distress: High | -0.004 (-0.024 to 0.015) | 0.653 | 0.009 (-0.019 to 0.038) | 0.501 | 0.002 (-0.011 to 0.016) | 0.739 | 0.001 (-0.024 to 0.025) | 0.961 |
| Minimally Sufficient Adjustment sets + peak BMI adjustment | | | | | | | | |
| Sex: Female | -0.031 (-0.041 to -0.021) | < 0.001 | 0.087 (0.072 to 0.101) | < 0.001 | -0.008 ( -0.016 to 0.000) | 0.052 | 0.027(0.014 to 0.039) | < 0.001 |
| Social Class: III | -0.002 (-0.017 to 0.014) | 0.821 | 0.009 ( -0.015 to 0.033) | 0.465 | 0.001 ( -0.010 to 0.011) | 0.915 | 0.009( -0.007 to 0.024) | 0.267 |
| Social Class: IV/V | -0.006 (-0.023 to 0.010) | 0.445 | 0.025 (0.002 to 0.048) | 0.036 | 0.000 ( -0.010 to 0.011) | 0.980 | 0.012( -0.007 to 0.030) | 0.204 |
| Birthweight (per 100g) | 0.001 (0.000 to 0.001) | 0.291 | 0.000 ( -0.002 to 0.001) | 0.852 | 0.000 ( -0.001 to 0.000) | 0.372 | 0.000( -0.001 to 0.001) | 0.755 |
| Breast Fed: No | -0.004 (-0.016 to 0.008) | 0.537 | 0.016 ( -0.006 to 0.038) | 0.139 | -0.001 ( -0.010 to 0.007) | 0.780 | 0.000( -0.013 to 0.013) | 0.951 |
| Housing Tenure: Rent/Other | -0.006 (-0.016 to 0.004) | 0.227 | 0.012 ( -0.007 to 0.031) | 0.202 | 0.003 ( -0.006 to 0.013) | 0.513 | 0.001( -0.012 to 0.014) | 0.910 |
| Childhood BMI (per 1kg/m^2^) | 0.001 (-0.002 to 0.003) | 0.586 | -0.003 ( -0.008 to 0.001) | 0.151 | 0.001 ( -0.001 to 0.003) | 0.254 | 0.000( -0.003 to 0.004) | 0.807 |
| Medical Conditions: Yes | -0.001 (-0.013 to 0.012) | 0.887 | 0.006 ( -0.008 to 0.020) | 0.398 | 0.000 ( -0.008 to 0.009) | 0.927 | -0.004( -0.017 to 0.010) | 0.588 |
| Psychological Distress: High | -0.005 (-0.021 to 0.012) | 0.557 | 0.010 ( -0.019 to 0.039) | 0.483 | 0.002 ( -0.011 to 0.015) | 0.787 | 0.001( -0.022 to 0.023) | 0.945 |

### Appendix A7: Early life drivers of weight change BCS70

**Appendix Table A9. Average marginal effects from multinomial logistic regression showing associations between potential early life drivers and eventual weight change group membership in BCS70**

**Appendix Table A9 Footnote:** For each individual driver of weight change, separate multinomial logistic regressions analysis were run, adjusting for a unique set of covariates, as deemed appropriate for the specific driver, using Directed Acyclic Graphs (DAGs). For each of the above models, the reference groups are as follows: Sex – male; Social Class - class I/II (managerial, professional, technical); Breast feeding – yes; Housing Tenure – own outright or with a mortgage; Medical Conditions – none; Psychological Distress – low.

### Appendix A8: Early life drivers of weight change NCDS

**Appendix Table A10. Average marginal effects from multinomial logistic regression showing associations between potential early life drivers and eventual weight change group membership in NCDS**

| NCDS Early Life Drivers of Weight Change | Obesity Weight Maintenance | | Obesity Weight Gain | | Obesity Weight Loss | | Obesity Weight Cycling | |
| --- | --- | --- | --- | --- | --- | --- | --- | --- |
|  | AME (95% CI) | P Value | AME (95% CI) | P Value | AME (95% CI) | P Value | AME (95% CI) | P Value |
| Minimally Sufficient Adjustment sets | | | | | | | | |
| Sex: Female | -0.016 (-0.025 to -0.008) | < 0.001 | 0.005 (-0.010 to 0.021) | 0.504 | 0.006 (0.001 to 0.012) | 0.016 | 0.001 (-0.009 to 0.012) | 0.819 |
| Social Class: III | 0.015 (0.001 to 0.028) | 0.030 | 0.051 (0.031 to 0.071) | < 0.001 | 0.004 (-0.005 to 0.014) | 0.329 | 0.010 (-0.006 to 0.025) | 0.220 |
| Social Class: IV/V | 0.021 (0.006 to 0.036) | 0.007 | 0.069 (0.043 to 0.095) | < 0.001 | 0.009 (-0.002 to 0.021) | 0.115 | 0.016 (-0.001 to 0.032) | 0.059 |
| Birthweight (per 100g) | 0.001 (0.000 to 0.002) | 0.009 | 0.001 (0.000 to 0.003) | 0.052 | 0.000 (0.000 to 0.001) | 0.562 | 0.000 (-0.001 to 0.001) | 0.663 |
| Breastfed: No | 0.008 (-0.002 to 0.018) | 0.117 | 0.007 (-0.010 to 0.024) | 0.399 | 0.003 (-0.004 to 0.011) | 0.372 | 0.009 (-0.003 to 0.022) | 0.138 |
| Housing Tenure: Rent/Other | 0.008 (-0.001 to 0.018) | 0.091 | 0.034 (0.015 to 0.052) | < 0.001 | 0.001 (-0.006 to 0.008) | 0.860 | 0.011 (-0.002 to 0.025) | 0.104 |
| Childhood BMI (per 1kg/m^2^) | 0.013 (0.012 to 0.015) | < 0.001 | 0.023 (0.020 to 0.026) | < 0.001 | 0.007 (0.006 to 0.008) | < 0.001 | 0.013 (0.011 to 0.015) | < 0.001 |
| Medical Conditions: Yes | -0.001 (-0.011 to 0.008) | 0.769 | 0.005 (-0.010 to 0.020) | 0.522 | -0.002 (-0.008 to 0.005) | 0.586 | -0.003 (-0.015 to 0.010) | 0.679 |
| Psychological Distress: High | 0.002 (-0.011 to 0.015) | 0.755 | 0.025 (0.001 to 0.048) | 0.041 | -0.001 (-0.010 to 0.008) | 0.832 | 0.003 (-0.012 to 0.018) | 0.730 |
| Minimally Sufficient Adjustment sets + peak BMI adjustment | | | | | | | | |
| Sex: Female | -0.018 (-0.026 to -0.010) | < 0.001 | 0.056 (0.041 to 0.071) | < 0.001 | 0.004 (-0.002 to 0.009) | 0.163 | 0.016 (0.006 to 0.027) | 0.003 |
| Social Class: III | 0.002 (-0.013 to 0.018) | 0.782 | 0.026 (0.006 to 0.047) | 0.012 | -0.003 (-0.013 to 0.008) | 0.634 | -0.005 (-0.021 to 0.012) | 0.568 |
| Social Class: IV/V | 0.003 (-0.014 to 0.019) | 0.739 | 0.031 (0.006 to 0.055) | 0.015 | 0.000 (-0.013 to 0.013) | 0.991 | -0.004 (-0.021 to 0.012) | 0.609 |
| Birthweight (per 100g) | 0.000 (-0.001 to 0.001) | 0.485 | 0.000 (-0.001 to 0.001) | 0.923 | 0.000 (-0.001 to 0.000) | 0.366 | 0.000 (-0.002 to 0.001) | 0.381 |
| Breast Fed: No | 0.002 (-0.007 to 0.010) | 0.692 | -0.003 (-0.019 to 0.012) | 0.669 | 0.000 (-0.007 to 0.007) | 0.924 | 0.004 (-0.008 to 0.015) | 0.524 |
| Housing Tenure: Rent/Other | -0.001 (-0.010 to 0.008) | 0.762 | 0.018 (0.001 to 0.035) | 0.039 | -0.005 (-0.012 to 0.002) | 0.160 | 0.003 (-0.010 to 0.016) | 0.623 |
| Childhood BMI (per 1kg/m^2^) | 0.000 (-0.001 to 0.001) | 0.988 | -0.004 (-0.008 to -0.001) | 0.016 | 0.001 (-0.001 to 0.002) | 0.290 | 0.000 (-0.002 to 0.002) | 0.874 |
| Medical Conditions: Yes | 0.001 (-0.008 to 0.010) | 0.812 | 0.010 (-0.005 to 0.024) | 0.184 | -0.001 (-0.007 to 0.006) | 0.815 | 0.000 (-0.012 to 0.012) | 0.962 |
| Psychological Distress: High | -0.003 (-0.014 to 0.008) | 0.604 | 0.018 (-0.002 to 0.038) | 0.078 | -0.004 (-0.012 to 0.004) | 0.346 | -0.001 (-0.015 to 0.013) | 0.881 |

**Appendix Table A10 Footnote:** For each individual driver of weight change, separate multinomial logistic regressions analysis was run, adjusting for a unique set of covariates, as deemed appropriate for the specific driver, using Directed Acyclic Graphs (DAGs). For each of the above models, the reference groups are as follows: Sex – male; Social Class - class I/II (managerial, professional, technical); Breast feeding – yes; Housing Tenure – own outright or with a mortgage; Medical Conditions – none; Psychological Distress – low.

### Appendix A9: Adult drivers of weight change in BCS70

| BCS70 Adult Driver of Weight Change | Obesity Weight Maintenance | | Obesity Weight Gain | | Obesity Weight Loss | | Obesity Weight Cycling | |
| --- | --- | --- | --- | --- | --- | --- | --- | --- |
|  | AME (95% CI) | P Value | AME (95% CI) | P Value | AME (95% CI) | P Value | AME (95% CI) | P Value |
| Minimally Sufficient Adjustment sets | | | | | | | | |
| HH Income: 4th | 0.001 (-0.015 to 0.017) | 0.934 | 0.001 (-0.024 to 0.025) | 0.962 | 0.000 (-0.010 to 0.010) | 0.994 | 0.007 (-0.015 to 0.030) | 0.511 |
| HH Income: 3rd | 0.007 (-0.009 to 0.023) | 0.420 | 0.032 (0.005 to 0.059) | 0.022 | 0.002 (-0.008 to 0.011) | 0.704 | 0.006 (-0.012 to 0.023) | 0.535 |
| HH Income: 2nd | 0.010 (-0.008 to 0.028) | 0.266 | 0.019 (-0.004 to 0.043) | 0.108 | 0.007 (-0.003 to 0.017) | 0.178 | 0.011 (-0.011 to 0.033) | 0.314 |
| HH Income: lowest | 0.005 (-0.014 to 0.024) | 0.266 | 0.050 (0.025 to 0.074) | < 0.001 | 0.006 (-0.004 to 0.016) | 0.260 | 0.011 (-0.010 to 0.031) | 0.314 |
| Education: No degree | 0.004 (-0.010 to 0.017) | 0.594 | 0.043 (0.024 to 0.061) | < 0.001 | 0.004 (-0.004 to 0.011) | 0.325 | 0.008 (-0.007 to 0.023) | 0.310 |
| LSI: Yes | 0.023 (0.010 to 0.036) | < 0.001 | 0.034 (0.013 to 0.055) | 0.002 | 0.010 (0.003 to 0.018) | 0.008 | 0.018 (0.005 to 0.031) | 0.009 |
| Smoking: Former | -0.002 (-0.018 to 0.000) | 0.810 | -0.012 (-0.034 to 0.010) | 0.287 | 0.005 (-0.003 to 0.014) | 0.214 | 0.000 (-0.015 to 0.000) | 0.953 |
| Smoking: Current | -0.024 (-0.037 to -0.011) | < 0.001 | -0.023 (-0.043 to -0.002) | 0.029 | 0.000 (-0.008 to 0.007) | 0.912 | -0.016 (-0.030 to -0.001) | 0.034 |
| Married/CP: Yes | 0.010 (-0.002 to 0.022) | 0.117 | -0.008 (-0.025 to 0.010) | 0.375 | 0.001 (-0.005 to 0.006) | 0.788 | 0.009 (-0.004 to 0.022) | 0.159 |
| Trying to lose weight: Yes | 0.066 (0.054 to 0.078) | < 0.001 | 0.129 (0.109 to 0.150) | < 0.001 | 0.018 (0.010 to 0.025) | < 0.001 | 0.060 (0.047 to 0.073) | < 0.001 |
| Minimally Sufficient Adjustment sets + peak BMI adjustment | | | | | | | | |
| HH Income: 4th | -0.001 (-0.017 to 0.014) | 0.857 | -0.003 (-0.026 to 0.021) | 0.805 | -0.001 (-0.011 to 0.010) | 0.876 | 0.006 (-0.016 to 0.028) | 0.597 |
| HH Income: 3rd | -0.002 (-0.018 to 0.013) | 0.761 | 0.015 (-0.009 to 0.039) | 0.208 | -0.001 (-0.011 to 0.009) | 0.865 | -0.004 (-0.022 to 0.014) | 0.674 |
| HH Income: 2nd | -0.001 (-0.018 to 0.016) | 0.882 | -0.004 (-0.026 to 0.018) | 0.738 | 0.003 (-0.007 to 0.014) | 0.532 | -0.002 (-0.023 to 0.019) | 0.849 |
| HH Income: lowest | -0.008 (-0.027 to 0.011) | 0.882 | 0.033 (0.009 to 0.056) | 0.008 | 0.001 (-0.009 to 0.012) | 0.843 | 0.000 (-0.020 to 0.020) | 0.849 |
| Education: No degree | -0.007 (-0.019 to 0.005) | 0.265 | 0.022 (0.003 to 0.040) | 0.021 | 0.001 (-0.006 to 0.008) | 0.825 | -0.004 (-0.019 to 0.011) | 0.575 |
| LSI: Yes | 0.002 (-0.011 to 0.014) | 0.794 | 0.006 (-0.013 to 0.024) | 0.545 | 0.003 (-0.004 to 0.009) | 0.449 | 0.000 (-0.011 to 0.012) | 0.933 |
| Smoking: Former | 0.001 (-0.013 to 0.000) | 0.860 | -0.009 (-0.030 to 0.011) | 0.359 | 0.006 (-0.002 to 0.014) | 0.136 | 0.002 (-0.012 to 0.000) | 0.775 |
| Smoking: Current | -0.008 (-0.020 to 0.005) | 0.225 | 0.002 (-0.018 to 0.022) | 0.869 | 0.005 (-0.002 to 0.013) | 0.162 | -0.002 (-0.017 to 0.014) | 0.844 |
| Married/CP: Yes | 0.004 (-0.007 to 0.015) | 0.443 | -0.019 (-0.035 to -0.003) | 0.020 | -0.001 (-0.006 to 0.004) | 0.674 | 0.003 (-0.010 to 0.017) | 0.609 |
| Trying to lose weight: Yes | 0.010 (-0.001 to 0.021) | 0.074 | 0.023 (0.006 to 0.040) | 0.010 | -0.001 (-0.007 to 0.005) | 0.739 | 0.000 (-0.015 to 0.014) | 0.961 |

**Appendix Table A11: Average marginal effects from multinomial logistic regression showing associations between potential adult drivers and eventual weight change group membership in BCS70**

**Appendix Table A11 Footnote:** The outcome for models or adult drivers of weight change, is based on the same weight change variable, but using BMI at 42 rather than peak BMI, to identify subsequent changes in weight. For each individual driver of weight change, separate multinomial logistic regressions analysis were run, adjusting for a unique set of covariates, as deemed appropriate for the specific driver, using Directed Acyclic Graphs (DAGs). For each of the above models, the reference groups are as follows: Income – highest; Education: - has a degree; Long Standing Illness (LSI) – none; Partnership Status – not legally married (or in a civil partnership); Trying to lose weight – no (measured in BCS70 only).

### Appendix A10: Adult drivers of weight change in NCDS

| NCDS Adult Driver of Weight Change | Obesity Weight Maintenance | | Obesity Weight Gain | | Obesity Weight Loss | | Obesity Weight Cycling | |
| --- | --- | --- | --- | --- | --- | --- | --- | --- |
|  | AME (95% CI) | P Value | AME (95% CI) | P Value | AME (95% CI) | P Value | AME (95% CI) | P Value |
| Minimally Sufficient Adjustment sets | | | | | | | | |
| HH Income: 4th | 0.000 (-0.016 to 0.015) | 0.974 | 0.010 (-0.019 to 0.038) | 0.301 | 0.000 (-0.011 to 0.010) | 0.603 | 0.001 (-0.018 to 0.020) | 0.938 |
| HH Income: 3rd | 0.002 (-0.014 to 0.017) | 0.805 | 0.015 (-0.016 to 0.045) | 0.504 | -0.002 (-0.013 to 0.009) | 0.939 | 0.006 (-0.011 to 0.023) | 0.472 |
| HH Income: 2nd | 0.000 (-0.014 to 0.014) | 0.969 | 0.025 (-0.005 to 0.055) | 0.338 | -0.002 (-0.011 to 0.007) | 0.692 | 0.005 (-0.011 to 0.020) | 0.569 |
| HH Income: lowest | -0.001 (-0.015 to 0.013) | 0.916 | 0.031 (0.000 to 0.061) | 0.098 | -0.002 (-0.012 to 0.007) | 0.615 | 0.014 (-0.005 to 0.033) | 0.145 |
| Education: No degree | 0.006 (-0.005 to 0.017) | 0.301 | 0.017 (-0.002 to 0.036) | 0.047 | 0.002 (-0.004 to 0.008) | 0.633 | 0.006 (-0.008 to 0.021) | 0.395 |
| LSI: Yes | 0.009 (-0.002 to 0.021) | 0.103 | 0.036 (0.017 to 0.055) | 0.079 | 0.004 (-0.002 to 0.009) | 0.491 | 0.010 (-0.002 to 0.023) | 0.110 |
| Smoking: Former | 0.002 (-0.009 to 0.013) | 0.713 | 0.007 (-0.013 to 0.028) | < 0.001 | 0.003 (-0.004 to 0.010) | 0.197 | 0.002 (-0.016 to 0.019) | 0.848 |
| Smoking: Current | -0.012 (-0.022 to -0.002) | 0.020 | -0.011 (-0.028 to 0.005) | 0.477 | -0.001 (-0.007 to 0.006) | 0.414 | -0.010 (-0.024 to 0.004) | 0.158 |
| Married/CP: Yes | 0.011 (0.002 to 0.020) | 0.017 | -0.005 (-0.024 to 0.015) | 0.185 | 0.002 (-0.003 to 0.007) | 0.817 | 0.005 (-0.006 to 0.016) | 0.411 |
| Minimally Sufficient Adjustment sets + peak BMI adjustment | | | | | | | | |
| HH Income: 4th | -0.001 (-0.015 to 0.013) | 0.904 | 0.008 (-0.018 to 0.034) | 0.535 | -0.001 (-0.011 to 0.010) | 0.779 | 0.000 (-0.019 to 0.019) | 0.982 |
| HH Income: 3rd | 0.001 (-0.014 to 0.017) | 0.869 | 0.010 (-0.018 to 0.038) | 0.462 | -0.002 (-0.013 to 0.008) | 0.907 | 0.004 (-0.012 to 0.021) | 0.591 |
| HH Income: 2nd | -0.001 (-0.014 to 0.012) | 0.868 | 0.022 (-0.006 to 0.049) | 0.122 | -0.003 (-0.012 to 0.006) | 0.657 | 0.003 (-0.012 to 0.018) | 0.694 |
| HH Income: lowest | -0.005 (-0.017 to 0.008) | 0.452 | 0.024 (-0.005 to 0.053) | 0.099 | -0.004 (-0.013 to 0.006) | 0.489 | 0.010 (-0.008 to 0.028) | 0.258 |
| Education: No degree | 0.001 (-0.010 to 0.012) | 0.813 | 0.005 (-0.013 to 0.023) | 0.592 | 0.001 (-0.006 to 0.007) | 0.415 | 0.001 (-0.013 to 0.015) | 0.922 |
| LSI: Yes | -0.002 (-0.011 to 0.008) | 0.750 | 0.009 (-0.007 to 0.025) | 0.262 | 0.000 (-0.005 to 0.005) | 0.801 | -0.001 (-0.013 to 0.011) | 0.864 |
| Smoking: Former | 0.001 (-0.009 to 0.011) | 0.786 | 0.001 (-0.017 to 0.019) | 0.935 | 0.003 (-0.004 to 0.009) | 0.936 | -0.001 (-0.017 to 0.015) | 0.931 |
| Smoking: Current | -0.004 (-0.014 to 0.005) | 0.381 | 0.009 (-0.007 to 0.025) | 0.264 | 0.002 (-0.005 to 0.008) | 0.435 | -0.002 (-0.016 to 0.012) | 0.757 |
| Married/CP: Yes | 0.008 (-0.001 to 0.017) | 0.068 | -0.020 (-0.038 to -0.003) | 0.026 | 0.001 (-0.003 to 0.006) | 0.605 | -0.001 (-0.011 to 0.010) | 0.873 |

**Appendix Table A12: Average marginal effects from multinomial logistic regression showing associations between potential adult drivers and eventual weight change group membership in NCDS**

**Appendix Table A12 Footnote:** The outcome for models or adult drivers of weight change, is based on the same weight change variable, but using BMI at 42 rather than peak BMI, to identify subsequent changes in weight. For each individual driver of weight change, separate multinomial logistic regressions analysis were run, adjusting for a unique set of covariates, as deemed appropriate for the specific driver, using Directed Acyclic Graphs (DAGs). For each of the above models, the reference groups are as follows: Income – highest; Education: - has a degree; Long Standing Illness (LSI) – none; Partnership Status – not legally married (or in a civil partnership); Trying to lose weight – no.
